# Effect of plasma treatment on the shear bond strength of ceramic and composite to human dental enamel

**DOI:** 10.1101/2025.07.14.25331489

**Authors:** Elnaz Shafigh, Mahdi sokhanvar, navid shafigh

## Abstract

**Background and Objective:** Cold atmospheric plasma (CAP) has the potential to decontaminate and disinfect carious lesions without the need for mechanical preparation. This study aimed to evaluate the effect of CAP on the bonding strength of composite and porcelain to enamel in an in vitro setting.

**Materials and Methods:** 44 sound premolar teeth extracted for orthodontic purposes were selected and randomly divided into two groups. The first group CAP treatment (CAP group), while the second group did not receive CAP treatment (NO CAP group). Each group was further subdivided into two subgroups: one prepared for bonding with E-max porcelain (E CAP and E NO CAP) and the other for bonding with composite resin (C CAP and C NO CAP).

In the composite subgroups, all samples were etched, rinsed and air-dried. They were then coated with a fifth-generation Kulzer bonding agent, light-cured, and subsequently bonded with composite disks (5 mm in diameter), which were light-cured again for 40 seconds.

In the E-max subgroups, the enamel surfaces were etched with HF acid, rinsed, and air-dried. They were then treated with Bisco silane, followed by application of the fifth-generation Kulzer bonding agent. The E-max disks (5 mm in diameter) were bonded using a self-adhesive dual-cure resin cement and cured for 2.5 minutes.

shear bond strength testing done using a Zwick/Rowell testing machine until debonding occurred. The failure mode (adhesive or cohesive) and debonding location were examined using a light microscope.

**Results:** Following plasma treatment, the median bond strength in the composite group increased from 88.7 to 92.6 MPa, while in the ceramic group, it increased from 174.5 to 187.4 MPa. Additionally, in the composite group, the number of cohesive failures increased from 3 in the NO CAP subgroup to 9 in the CAP subgroup. Similarly, in the ceramic group, the number of cohesive failures increased from 1 to 3 after CAP treatment.

**Conclusion:** the effect of CAP on increasing cohesive failures in ceramics was not significant, whereas in the composite group, this effect was statistically significant.

## Introduction

The oral cavity is recognized as a complex microbial habitat, home to more than 700 microbial species. These microorganisms, as part of the natural flora, interact harmoniously with the human immune system.

Periodontal diseases and dental caries are among the most prevalent challenges in dentistry. Dental caries, a leading cause of toothache, may occur in the crown or root of the tooth. This phenomenon is defined as the localized destruction of dental tissue due to acid production by bacteria. ^1^The acids produced lead to a decrease in the local pH of the tooth below the critical level, resulting in demineralization of the dental tissue .^2^ If the loss of calcium, phosphate, and carbonate ions from the tooth structure continues, a carious lesion will eventually form .^3^

Global studies, including a 2017 report by The Lancet, have shown that among 328 diseases, permanent dental caries ranks first in prevalence and second in incidence. It is estimated that approximately 4.4 billion people worldwide suffer from this condition .^4^ Dental caries can affect the enamel, cementum, or dentin. The process often begins with small areas of demineralization in the enamel, which, in the early stages, may be halted or reversed by the absorption of calcium, phosphate, and fluoride ions. However, in most cases, without appropriate treatment, the demineralization process extends to the dentin and pulp, ultimately leading to the loss of tooth vitality .^5^ To treat damaged teeth, dentists recommend removing the decayed areas and filling the cavity with suitable restorative materials .^6^

Although restorative treatments, despite their limitations and the potential need for re-restoration, remain the preferred approach in many countries, ^7^ in some regions, such as Scandinavian countries, preventive methods have gradually replaced restorative treatments .^8^

Today, polymeric composites are widely used as dental restorative materials due to their superior properties, including biocompatibility, aesthetics, antibacterial properties, and non-toxicity. Although it took considerable time for resin composites to be recognized as suitable materials for anterior and posterior restorations, the development of these materials has led to improvements in their physical, mechanical, and thermal properties. During the era when amalgam was considered the gold standard for posterior restorations, the first generation of dental composites faced challenges such as low wear resistance and poor mechanical strength.

Recent advancements, including the optimization of the resin phase, reduction of polymerization shrinkage, enhancement of fracture toughness, and the use of smaller filler particles with different morphologies, have increased the longevity of dental composites and reduced restoration failures. Additionally, approaches such as incorporating antibacterial and self-healing properties are being explored to enhance the performance and durability of these materials .^9^

E-max ceramic restorations represent a significant advancement in dentistry, particularly in the field of dental prosthetics. These restorations are made from a high-quality glass-ceramic material known as lithium disilicate, renowned for its exceptional aesthetics and high strength. The use of E-max enables dentists to create highly durable and aesthetically pleasing restorations that closely resemble the natural appearance of teeth. This innovative material is especially beneficial for patients seeking both functional and cosmetic improvements in their dental health, providing a reliable solution for various restorative needs, including crowns, veneers, and dental bridges. The introduction of E-max ceramic restorations has revolutionized aesthetic dentistry, delivering superior outcomes for patients .^10^

### Enamel Etching and Bonding Agents

Etching enamel with acid is an effective method for increasing the adhesion of resin-based restorative materials. The first generation of acid-etching systems was introduced in the mid-1970s. With advancements in dental materials, various formulations and application protocols for acid etchants have been developed. Today, phosphoric acid at concentrations between 32% and 40% is widely used for bonding resin materials to enamel surfaces.

Laboratory studies on the effects of different phosphoric acid concentrations on enamel have shown that concentrations below 30% are insufficient to create the necessary enamel dissolution for improved resin adhesion. Similarly, concentrations above 50% do not provide an optimal surface morphology for bonding .^11^

The primary objectives of the etching process include chemical cleaning of the enamel surface, improving its wettability, increasing the available surface area for contact with polymeric materials, and creating micro porosities that facilitate the formation of resin tags .^12^

Bonding agents play a crucial role in dentistry by ensuring the adhesion between restorative materials and the natural tooth structure. These materials are specifically formulated to create strong bonds, ensuring that various restorative materials, such as composite resins, effectively adhere to enamel and dentin surfaces. The effectiveness of bonding agents is critical for the longevity and durability of dental restorations, as they help prevent issues such as microleakage and secondary caries. Additionally, bonding agents contribute to the aesthetic outcomes of dental procedures by allowing seamless integration of restorative materials with the natural appearance of teeth. Their use is fundamental in various dental procedures, including restorations, veneers, and crowns, underscoring their importance in modern dentistry.

To fulfill these challenging tasks, various chemical compositions and techniques are continuously developed and integrated into bonding agents. The incorporation of monomers resistant to hydrolytic degradation and functional monomers that enhance micromechanical retention and increase chemical interactions between adhesive resin materials and different substrates has improved the performance of bonding agents. The current trend involves incorporating bioactive molecules into adhesive materials to enhance mechanical properties and prevent enzymatic degradation of the dental substrate, thereby ensuring the durability of resin-dentin bonds. Additionally, alternative etching materials and techniques have been developed to address the limitations of phosphoric acid etching of dentin. Overall, contemporary adhesive dentistry is experiencing a dynamic period, with advancements aimed at simplifying and improving the reliability of bonding processes. However, simplification and innovation should not come at the cost of reduced material quality .^13^ in other hand failure interactions are practically done mainly through creating a tiny crack as the source of the damage and its growth and progression. For a crack to advance in a material, the concentration stress at the crack tip must exceed the tensile strength at that position and the etching ability of plasma can reduce the tension and crack grows. The possible failure of dental ceramics is due to the accumulation and growth of micro damage. Brittle fracture is the most critical failure mechanism in dental ceramics. No plastic deformation is observed during this sudden failure. The brittle fracture occurs with cracks around the defect^14^

### Plasma in Dentistry

Plasma, recognized as the fourth state of matter, was first identified in 1879 by British physicist Sir William Crookes and was named by American chemist Irving Langmuir in 1929. Plasma consists of charged and excited particles and is the most common state of matter in the universe, comprising over 99% of the visible cosmos. This state occurs when electrons separate from atoms or molecules, transforming the material into plasma. If energy is removed from the system, electrons recombine with heavier particles, reverting the material to a gaseous state. Unlike ordinary gases, plasma can exist over a wide temperature range without changing state.

Plasma can be generated under various conditions, including atmospheric pressure. In recent years, atmospheric plasmas have become attractive tools for material processing due to their usability in open environments. However, generating plasma under these conditions requires a high electric field, and without specific measures, plasma may overheat, potentially damaging sensitive materials or living tissues .^15^

Cold atmospheric plasma (CAP) has been introduced for applications involving living tissues. This plasma type operates at temperatures below 40°C, producing reactive species that include charged particles, radiation, and reactive oxygen species. CAP has gained attention for its antimicrobial properties, cellular modification capabilities, and potential in cancer cell destruction, making it a promising approach in dentistry and oncology .^16^ The ability of cold plasma to eradicate bacteria without harming surrounding tissues makes it a viable alternative to conventional methods .^17^

## Method materials

A number of premolar teeth with healthy crowns were selected for study. The enamel of the labial surface of the teeth was cleaned to remove any residual blood and tissue using a rubber cup and pumice, then washed and dried. The crowns of the teeth were examined under a light microscope by an operator for integrity, with special attention given to cracks and decay. Problematic teeth were excluded from the study. To facilitate handling, the teeth were mounted in putty. (Figure 3-1)

Subsequently, the enamel rods of the labial surface were freshened using a square blue polishing disk measuring 5×5 mm. After each set of five teeth was polished, the disk was replaced. A total of 44 premolar teeth, whose enamel rods had been polished with the disk, were prepared and randomly divided into two groups: CAP and CAP NO. Each group was further subdivided into two subgroups: C (composite) and E (ceramic). The samples were disinfected using an ultrasonic device (YAXUN China, Beijing, A2000YX) and then placed in distilled water at room temperature.

(Figure 3-2) A wooden piece with a diameter of 5 mm was selected, and circular cuts with a diameter of 5 mm were made in it using a laser cutting device (Figure 3-3). Using the cylinder method, 22 wax discs with a 5 mm diameter were fabricated, then scanned with a Shining D3 scanner (Hangzhou, China). The samples were 3D-printed using a K8 Sonic Phrozen printer (Hsinchu, Taiwan) into lithium disilicate Max-E discs (Figure 3-4). Composite material LX Palfique (Japan, Tokoyama) and machined wooden pieces were also used to create 22 composite 5 mm diameter discs (Figure 3-5).

The samples from both subgroups of the CAP group were removed from the distilled water and dried. The surface of the samples was then exposed to cold plasma (kv 16) for 30 seconds (Figure 3-6). In this study, a DBD probe was used to produce plasma for sterilization purposes (Dento Panel, Fanavaran Sepid Jamegan, Tehran, Iran) (Figures 3-7 and 3-8). The probe was placed as close as possible to the samples without direct contact. The DBD probe, utilizing a glass ampoule and neon gas, facilitates the transmission of plasma on the glass surface, ionizing air molecules and generating numerous active species.

### Application of Acid

The samples from the C subgroup of both groups (CAP and CAP NO) were removed from the distilled water and dried. The surfaces were then exposed for 30 seconds to a 37% phosphoric acid gel (Parlaco, Exetch, Iran, Tehran) (Figure 3-9). After the acid application, the samples were rinsed with a spray for 30 seconds and dried for 20 seconds.

The samples from the E subgroup of both groups (CAP and CAP NO) were also removed from the distilled water and dried. Their surfaces were exposed to a 9.6% hydrofluoric acid gel (Dentonext, Iran, Tehran) for 90 seconds. After the acid application, the samples were rinsed for 30 seconds with a spray and dried for 20 seconds.

### Bonding Procedure

The samples from the C subgroup of both groups (CAP and CAP NO) were exposed to a fifth-generation bonding agent (Kulzer, Germany, Group Chemical Mitsuai) using a microbrush, then air-dried for 10 seconds with a Malim air blower and cured for 20 seconds with an Elite Cure light-curing unit (Figure 3-10).

The samples from the E subgroup of both groups (CAP and CAP NO) were exposed to a silane treatment (Bisco, A.S.U., BISCO) for 30 seconds, then allowed to rest. Subsequently, the samples were treated with a fifth-generation bonding agent (Kulzer, Germany, Group Chemical Mitsuai), air-dried for 10 seconds, and a dual-cure resin cement, Cem-U (South Korea, Vericom), was applied on the uncured Kulzer bonding agent (Figure 3-12).

### Bonding Process for Both Subgroups

The samples from the C subgroup of both groups (CAP and CAP NO) were placed in contact with composite discs and exposed to the Elite Cure light for 30 seconds. The samples from the E subgroup of both groups (CAP and CAP NO), while still covered with uncured fifth-generation bonding agent and resin cement, were placed in contact with ceramic discs and then cured for 150 seconds.

To ensure precision in the measurement of the bond strength and improve ease of handling during testing, all the samples were mounted in pink acrylic using a clamp device. The shear bond strength was measured using a Roell/Zwick device at the Shahid Beheshti University of Dentistry (Figure 3-14).

### Microscopic Examination

The samples were examined under a light microscope (Euromex model Edublue, magnification 2x) to assess the debonding sites and determine the type of debonding (cohesive or adhesive) (Figure 3-16).

### Findings

By examining the numerical data obtained from the Zwick/Roell device regarding the maximum shear force that the bond can withstand before separation, the results for the two groups, CAP and NO CAP, and the two subgroups, C and E, are presented in Table 1.

**Table 1:**
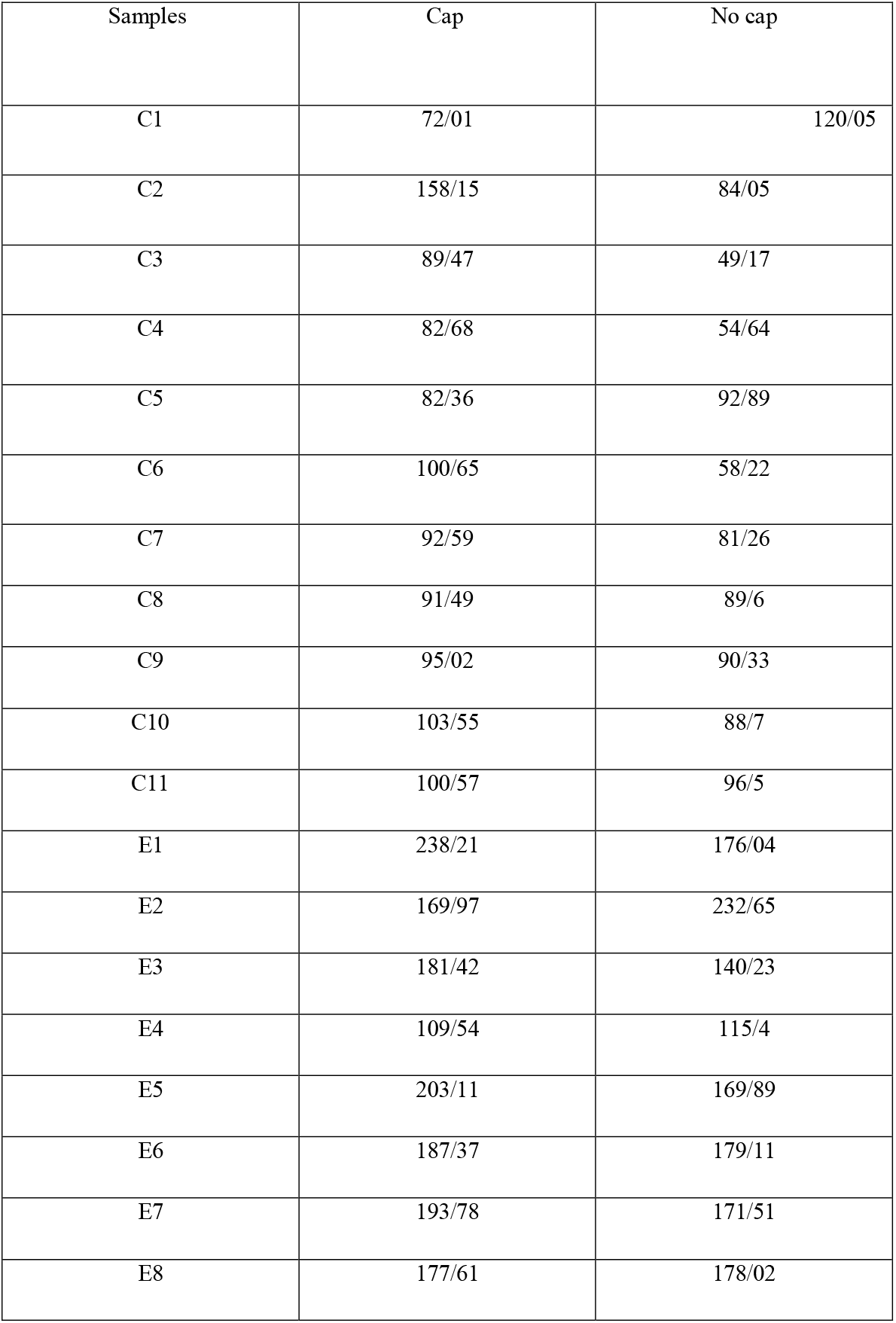
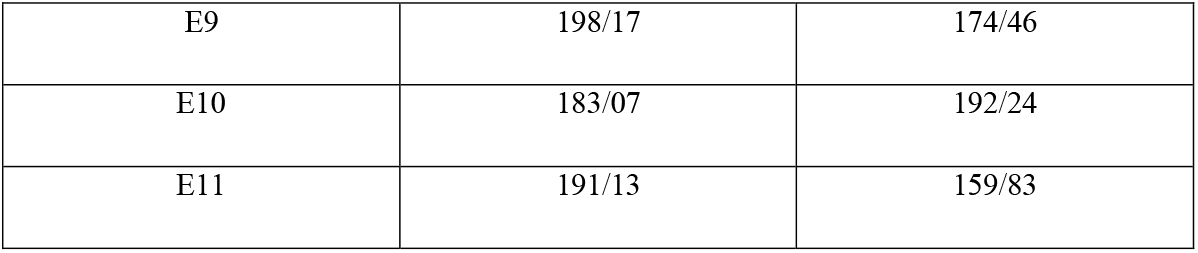
Shear bond strength of samples.

Due to the non-normal distribution of the samples, we used the median instead of the mean to symbolically represent the variation in the tested samples. The results are presented in Figure 1. Under the conditions of this experiment, the median of the composite subgroup increased from 88.7 to 92.6 after cold plasma treatment, while the median of the ceramic subgroup increased from 174.5 to 187.4 following the same treatment.

**Figure 1:**
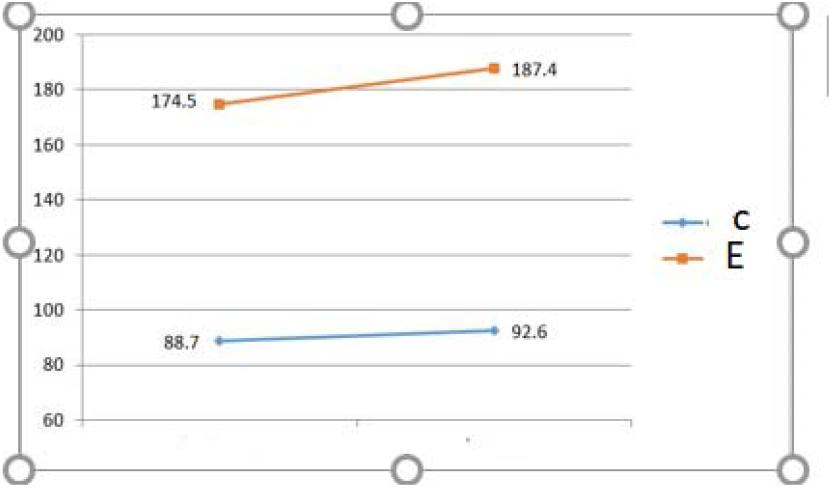
effect of plasma on median SBS of samples Upper one representing ceramic and the lower one, composite.

The analysis of debonding type (cohesive or adhesive) using an optical microscope provided data detailed in Table 2. It is noteworthy that all cohesive failures in the ceramic group were of the dental cohesive type, where a portion of the tooth fractured during debonding. In contrast, in the composite cohesive group, only two samples exhibited cohesive failure, both of which belonged to the plasma-treated group.

**Table 2:** cohesive-adhesive bonding results.

| Failure mode of ceramics | cap | nocap |
| --- | --- | --- |
| cohesive | 3 | 1 |
| Adhesive | 8 | 10 |

| Failure mode of composites | cap | nocap |
| --- | --- | --- |
| cohesive | 9 | 3 |
| Adhesive | 2 | 8 |

Data analysis using SPSS indicated a non-normal distribution. Therefore, to determine the statistical significance of changes in shear bond strength, the non-parametric Mann-Whitney U test was employed with a significance threshold of less than 0.05. Due to the non-normal distribution, median values were used instead of means in the shear bond strength variation charts. Standard deviation, median, and interquartile range (IQR) were calculated separately for each group. Given the non-normal distribution, the Mann-Whitney U test was applied to assess the statistical significance of the increase in bonding strength of ceramic and composite to dentin and enamel after cold plasma treatment, with a significance level of less than 0.05.

### Data analyses

By analyzing the data obtained from the Zwick/Roell device regarding the maximum shear force that the bond can withstand before debonding, the results for the two groups, CAP and NO CAP, and their subgroups, C and E, are presented in Table 1.

Due to the non-normal distribution of the samples, the median was used instead of the mean to symbolically represent variations in the tested samples. The results are shown in Figure 1. Under the conditions of this study, the median of the composite subgroup increased from 88.7 to 92.6 after cold plasma treatment, while the median of the ceramic subgroup increased from 174.5 to 187.4 after treatment.

The type of bond failure (cohesive or adhesive) was examined using an optical microscope, and the corresponding data are summarized in Table 2. It is noteworthy that all cohesive failures in the ceramic group were of the dental cohesive type, where a portion of the tooth fractured during debonding. In contrast, in the composite cohesive group, only two samples exhibited cohesive failure, both of which belonged to the plasma-treated group.

Data analysis using SPSS indicated a non-normal distribution. Therefore, to determine the statistical significance of changes in shear bond strength, the non-parametric Mann-Whitney U test was employed with a significance threshold of less than 0.05. Due to the non-normal distribution, median values were used instead of means in the shear bond strength variation charts. Standard deviation, median, and interquartile range (IQR) were calculated separately for each group and presented in Table 3. Given the non-normal distribution, the Mann-Whitney U test was applied to assess the statistical significance of the increase in bonding strength of ceramic and composite to dentin and enamel after cold plasma treatment, with a significance level of less than 0.05.

The results of this test, with a significance level of less than 0.05, indicated that although plasma treatment improved the bonding strength of ceramic and composite to enamel and dentin, this effect was not statistically significant. The Chi-square test, with a significance level of less than 0.05, showed that plasma treatment had a statistically significant effect on the cohesive failure of the composite bonding group, but not on the ceramic group. Plasma treatment did not significantly influence the cohesive failure of the ceramic bond to dentin and enamel.

## Discussion

Shafigh (2024) ^18^conducted a study on 42 teeth to assess the impact of cold plasma treatment on enamel resistance to acid etching. By examining samples under a scanning electron microscope, the mean demineralization depth was measured at 11.37 microns in the plasma-treated group and 7.65 microns in the untreated group. This study demonstrated that cold plasma treatment significantly increased demineralization depth (P<0.05). The findings align with the present study, as the increased demineralization depth led to enhanced shear bond strength and cohesive bond formation, though this increase in shear bond strength was not statistically significant. This discrepancy may be attributed to three factors:

1. The present study had more limiting factors than Shafigh’s study.
2. Demineralization depth is assessed on a micrometer scale, whereas significant changes in shear bond strength depend on multiple variables beyond just increased demineralization depth.
3. The current study had a sample size similar to the previous study but investigated two variables simultaneously, necessitating larger-scale studies for more definitive conclusions (28).

Kostadinov (2024)^19^ aimed to evaluate the effect of cold plasma pretreatment on the bond strength of two-piece hybrid ceramic crowns in implant prosthetics as an alternative to conventional methods. By testing eight groups, he sought the best combination for bonding these prostheses. His findings indicated that the highest bond strength occurred with the conventional combination of HF acid etching and silane treatment. Among the non-HF groups, the silane and plasma combination exhibited the highest bond strength, though it was 24% lower than the conventional method. These results contradict the present study, which found that plasma treatment alongside silane and HF increased bond strength, albeit not significantly. However, Kostadinov et al. concluded that two-factor interactions between silane and HF were not statistically significant, whereas three-factor interactions involving HF, silane, and plasma were significant at P<0.0001. This finding aligns with the present study, emphasizing the need for further research with larger sample sizes to obtain more conclusive results .^19^

Valizadeh (2021) ^20^examined the effect of cold plasma treatment on bovine incisors and changes in shear bond strength using three bonding agents: G-Premio Bond, Clearfil SE Bond, and Adper Single Bond. The study revealed that plasma treatment with G-Premio Bond significantly increased shear bond strength, while no significant difference was observed in the other groups. These results are in line with the present study, as the use of the fifth-generation Kulzer bonding agent in this study led to an increase in median shear bond strength in the composite group, though the increase was not statistically significant. Valizadeh’s study suggests that adjusting test conditions and the choice of bonding agent might establish a statistically significant relationship between plasma treatment and composite shear bond strength in laboratory conditions 20

Banz et al. (2021) investigated the shear bond strength of zirconia and composite discs using various methods, including plasma treatment in addition to sandblasting and Scotchbond Universal application. Their results varied: in the short term, the plasma group did not exhibit a positive effect, and the sandblasting and Scotchbond Universal group had the highest median SBS.^21^ However, in the long term, the plasma group demonstrated higher SBS. This finding aligns with the present study and highlights the necessity of repeating the current study using an artificial aging method, which could contribute to statistically significant results.^22^

Hirata et al. (2016) examined the effect of atmospheric plasma on microtensile bond strength to dentin with two adhesive systems (etch-and-rinse and self-etch) after one week and one year of water storage. In the one-week evaluation, plasma application after acid etching significantly increased bond strength and the percentage of cohesive failures. In contrast, for groups where plasma was applied before acid etching, no significant differences in bond strength or failure patterns were observed compared to control groups. The absence of bond strength changes for plasma-treated groups before acid etching contradicts the present study .^23^

Hirata et al. (2015) assessed the effect of atmospheric plasma on microtensile bond strength to dentin with self-etch adhesives after one year of water storage. They found that plasma treatment increased bond strength for one tested adhesive, which could be attributed to changes in dentin surface properties and increased demineralization depth, aligning with the present study .^24^

Teixeira et al. (2015) examined the effect of atmospheric plasma on the mechanical properties of enamel and the bond strength of sealants. Their findings indicated that plasma increased surface wettability and bond strength between enamel and sealants, potentially serving as an alternative to conventional acid etching methods or as an adjunct to self-etch sealants, supporting the results of the present study.

Dental ceramics are well known for restoring the function and aesthetic of lost or damaged teeth. Understanding these materials’ mechanical and aesthetic properties can make a suitable choice for those materials. The longevity of dental ceramics depends on several factors, including manufacturing method, clinical process, and the oral cavity’s aqueous environment. Failure mechanisms in restorative ceramics are complex and a combination of several factors. Different microstructures in the crystalline phase will involve the propagation of cracks and eventually the fatigue of ceramic materials. Complementary studies needed to understand the effect of crystalline phases on the crack grows and bond strength of these materials. .^25^

## Conclusion

The present study demonstrated that under the given conditions, atmospheric cold plasma treatment increased the bond strength of composite and ceramic to dentin and enamel, though this increase was not statistically significant. However, this method significantly enhanced the cohesive bond of composite to enamel and dentin.

## Data Availability

All data produced in the present work are contained in the manuscript

## Acknowledgments

The authors sincerely thank all professors and experts who contributed to this research. Special appreciation is extended to the esteemed faculty of the Aja School of Dentistry.

## Declaration

### Ethics approval and consent to participate

The protocol of the present research was approved by the university ethics committee with Ethical Approval Number IR.AJAUMS.REC.1403.141 Consent for publication: all authors have consent for publication and declare this in the consent form.

### Availability of data and materials

The databases and all of the findings of this study are available on request from the corresponding author. The data are not publicly available because of ethical restrictions.

## AUTHOR CONTRIBUTIONS

All authors contributed to the conception, design, data acquisition and interpretation, and statistical analysis and drafted and critically revised the manuscript. All authors gave their final approval and agree to be accountable for all aspects of the work.

## Clinical trial number

not applicable.

